# Platform-Specific Safety of Pulsed Field Ablation for Atrial Fibrillation: A MAUDE Analysis

**DOI:** 10.64898/2026.07.13.26358008

**Authors:** Ahsan Ullah, Jose Fossas Espinosa, Luka Petrovic, Emad F. Aziz

## Abstract

**Background:** Three pulsed-field ablation (PFA) systems are FDA-approved for atrial fibrillation (AF), yet whether their safety profiles differ from each other and from radiofrequency (RF) ablation has not been systematically characterized using real-world adverse event data. We compared platform-specific complication profiles across three FDA-approved PFA systems and contemporary RF catheters in MAUDE.

**Methods:** We analyzed 2,262 manually adjudicated MAUDE adverse event reports (760 PFA, 1,502 RF) through July 2025. Neurologic events underwent independent adjudication into five tiers by three auditors. Disproportionality was assessed using Reporting Odds Ratios (ROR) with Benjamini-Hochberg (BH) correction.

**Results:** Pooled PFA had significantly lower BH-adjusted ROR for tamponade (0.52, 95% CI 0.41-0.67) and esophageal injury (0.09, 0.01-0.66), consistent with a tissue-selective reporting profile across platforms. Platform-level analysis, however, revealed substantial heterogeneity: the stroke signal was driven by Varipulse (ROR 16.41, 8.61-31.28) and was not observed with Farapulse (ROR 1.26, NS). Pooled PFA had higher ROR for imaging-confirmed stroke (3.84, 2.27-6.49) and arrhythmia (2.57, 1.91-3.45). Coronary vasospasm (24 vs. 0 events) and hemolysis (15 vs. 1 events) were PFA-specific. Composite serious adverse events were similar. In a pre-specified extension period analysis (August-December 2025), the pooled PFA stroke signal attenuated to non-significance (ROR 1.60, 0.89-2.85), consistent with notoriety bias following the FDA Safety Communication.

**Conclusions:** PFA adverse-event reporting shows substantial platform heterogeneity across approved systems. Varipulse was associated with a disproportionate neurologic reporting signal, while all PFA platforms show tissue-selective reporting patterns relative to RF. These findings support platform-aware clinical decision-making and post-market surveillance.

## Introduction

Radiofrequency (RF) ablation for atrial fibrillation is a mature procedure. While modern iterations have reduced complications, RF ablation carries an established risk profile including atrioesophageal fistula, phrenic nerve injury, and pulmonary vein stenosis resulting from collateral thermal injury to adjacent structures, as well as cardiac tamponade from mechanical perforation or steam pop-related tissue disruption.

Pulsed field ablation (PFA) has emerged as a transformative alternative, leveraging irreversible electroporation, a non-thermal mechanism that selectively disrupts cardiomyocyte membranes while largely sparing adjacent tissue [2]. Three distinct PFA systems have received FDA approval for AF ablation: the Farawave pentaspline multielectrode catheter (Farapulse, Boston Scientific), the VARIPULSE variable-loop circular catheter (VLCC; Biosense Webster/Johnson & Johnson MedTech), and the Sphere-9 lattice-tip focal catheter with hybrid PFA/RF capability (Affera Mapping and Ablation System, Medtronic). These platforms differ substantially in electrode geometry, pulse parameters, energy delivery architecture, and irrigation design. Despite the rapid global adoption of PFA-with hundreds of thousands of procedures performed worldwide [24]-pivotal clinical trials enrolled highly selected populations with limited statistical power to detect rare complications. The ADVENT trial (Farapulse, n=305 PFA) [21] and the VARIPULSE IDE trial (n=118) [13] were powered for efficacy, not safety, and neither was designed to detect complications occurring at rates below 1-2% [3, 4]. The substantial differences in device architecture across approved PFA systems, combined with incomplete safety characterization in pivotal trials, highlight the need for comprehensive, platform-differentiated post-market safety analysis. The recent Varipulse voluntary pause (January 2025) and subsequent FDA Safety Communication (July 2025) regarding neurological events underscore the timeliness of this question.

The FDA MAUDE database is the principal publicly available post-market adverse event surveillance system for cleared medical devices in the United States [5]. MAUDE captures mandatory reports from manufacturers and healthcare facilities, providing a signal-detection capability that complements controlled-trial evidence. A recent MAUDE analysis by Cho et al. evaluated pooled PFA versus RF complications but was not designed to stratify adverse events by individual platform or to adjudicate neurologic events [6]. Building on this work, we performed a platform-level analysis of all three FDA-approved PFA systems and contemporary RF catheters, incorporating structured neurologic adjudication across five prespecified tiers, to characterize platform-specific adverse-event reporting profiles and identify disproportionate safety signals (Figure 5).

## Methods

### Study Design and Data Source

We performed a retrospective cross-sectional analysis of adverse event reports submitted to the FDA MAUDE database. This study used publicly available, de-identified data and was exempt from institutional review board oversight. The report conformed to the STROBE checklist for observational studies [1].

### Device Selection and Search Strategy

Five catheter-based ablation platforms were included: Farapulse (Boston Scientific); VARIPULSE (Biosense Webster); Affera (Medtronic); ThermoCool STSF (Biosense Webster); and QDOT Micro (Biosense Webster). The Medtronic PulseSelect system, which received FDA clearance in December 2023, was excluded because it employs a linear multi-electrode array designed for point-by-point ablation, a fundamentally different catheter architecture from the single-shot designs of the three included PFA platforms, and had limited U.S. commercial adoption during the study period with insufficient MAUDE report volume for platform-level disproportionality analysis. MAUDE was queried using manufacturer-specific brand-name terms in the device Brand Name field, filtered by manufacturer. The following search strings were applied: Farapulse/Farawave/Farastar (manufacturer: Boston Scientific); Varipulse/TruPulse (manufacturer: Biosense Webster); Affera/Sphere-9 (manufacturer: Medtronic); ThermoCool SmartTouch SurroundFlow/STSF (manufacturer: Biosense Webster); and QDOT Micro/QDOT (manufacturer: Biosense Webster). This brand-name approach yields a narrower, clinically focused cohort than a Product Code-based search, which captures all reports filed under a device classification code regardless of clinical context (including device malfunctions without patient contact, non-AF procedures, and international filings). The search window extended from each PFA device’s initial U.S. FDA clearance through July 31, 2025 (Farapulse: Feb 2024; Varipulse: Sep 2024; Affera: Jan 2024). Reports were indexed by MAUDE report receipt date (Date Received), which reflects the date the report was received by FDA rather than the date the adverse event occurred. For the RF comparators (QDOT and STSF), the analysis window began January 2024 to establish a contemporary post-PFA-era comparison period, although both devices received FDA clearance prior to this date. Cryoballoon ablation was excluded as the objective was to evaluate PFA against contemporary RF during the post-PFA-approval window. Report types included Injury and Death; Malfunction reports were included when narratives described patient harm.

### Inclusion and Exclusion Criteria

Reports were included if they identified a specific device and documented an adverse event occurring during or after AF ablation in an adult patient. Reports describing non-AF ablation procedures were excluded unless AF ablation was concurrently documented. Manufacturer follow-up reports were collapsed with the originating report using the FDA event key and report number to prevent double-counting. Reports identified as literature-referenced submissions and MAUDE filings citing a published study rather than a directly observed adverse event, as indicated by characteristic narrative phrasing, were excluded from the primary analysis and retained for a pre-specified sensitivity analysis.

### Complication Categorization and Adjudication

Adverse events were classified into nine complication domains consistent with the HRS/EHRA consensus definitions [7]: (1) arrhythmia (ventricular tachycardia/fibrillation, ablation-induced organized atrial arrhythmia, cardiac arrest, and high-grade AV block); (2) cardiac perforation/tamponade (including pericardial effusion requiring intervention); (3) neurologic event (stroke, TIA, and cerebral embolism); (4) death; (5) hemolysis (laboratory-confirmed or clinically significant hemoglobin-related sequelae); (6) pulmonary vein stenosis; (7) phrenic nerve injury; (8) esophageal injury (thermal, mechanical, or fistula formation); and (9) coronary vasospasm. A single report could contribute to multiple categories.

Given the clinical gravity of neurologic events, all reports flagged for stroke or TIA underwent a rigorous, structured adjudication process by three auditors (A.U., J.F.E., E.A.). Events were classified into a five-tier system: (1) **Strict (Imaging-Confirmed):** reports explicitly documenting acute stroke confirmed by CT or MRI; (2) **Moderate (+Silent Cerebral Lesion):** reports noting new, procedure-related silent cerebral lesions on imaging without clinical deficit; (3) **Broad (+Unconfirmed):** reports describing clinical stroke symptoms without documented imaging confirmation; (4) **Most Inclusive (+TIA):** reports describing transient ischemic attack symptoms; and (5) **Original (Pre-Adjudication):** all MAUDE reports with any neurology-related flag prior to adjudication (Supplemental Table S2). The **Strict (Imaging-Confirmed)** tier was used for the primary analysis to ensure the highest specificity. Disagreements were resolved by majority vote among the three auditors. Inter-auditor agreement was assessed across all 156 reports flagged for neurologic events (Fleiss’ kappa = 0.92); fewer than 8% of cases required majority-vote resolution.

Non-neurologic domains were categorized using a standardized keyword algorithm applied to MAUDE narrative fields, followed by comprehensive manual review that corrected false positives (e.g., transesophageal echocardiography coded as esophageal injury, negation-triggered flags) and false negatives (e.g., uncaptured cardiac arrest events). Non-neurologic categories were reviewed by a single auditor (A.U.) with verification by a second auditor (E.A.). Reports with ambiguous or unclassifiable narratives (e.g., insufficient clinical detail to assign a specific complication domain, or death without a documented proximate cause) were retained in the platform’s total report count but were not assigned to any specific complication domain. No reports were excluded from the analysis solely on the basis of narrative ambiguity. A single report could be classified into multiple complication domains when the narrative documented more than one distinct adverse event.

### Statistical Analysis

Adverse event counts and proportions were computed descriptively for each platform and complication domain. Disproportionality was quantified using the Reporting Odds Ratio (ROR), a standard and validated pharmacovigilance signal-detection method for spontaneous reporting databases [8]. All statistical analyses were performed using Python 3.11 (Python Software Foundation).

RORs and 95% confidence intervals (CIs) were calculated using the Woolf logit method. The Haldane-Anscombe correction (adding 0.5 to all cells) was applied to address zero-event cells and small platform denominators. Statistical significance for all comparisons was assessed using the two-tailed Fisher’s exact test, which is robust for small sample sizes.

To control the false discovery rate across the seven formally compared complication domains, raw p-values were adjusted using the Benjamini-Hochberg (BH) step-up procedure with a predefined false discovery rate threshold of q<0.05. PFA-specific mechanisms with ≤1 RF event (hemolysis, coronary vasospasm) are reported descriptively (Supplemental Table S1). A clinically meaningful disproportionality signal was prespecified as ROR >2.0, lower 95% CI >1.0, and ≥3 reported events. A composite serious adverse event endpoint was defined to include death, stroke/TIA, pericardial tamponade or perforation, and esophageal injury.

### Sensitivity Analysis

Primary analyses were repeated including the 52 literature-referenced reports excluded from the primary cohort. False positives identified during manual QC were excluded.

### Post-IFU Temporal and Extension Period Sensitivity Analyses

To evaluate signal durability following the Varipulse IFU revision, we analyzed Varipulse and RF reports received between August 2025 and April 2026 (Supplemental Table S10). To evaluate notoriety bias following the July 2025 FDA Safety Communication, we performed a pre-specified extension period analysis comparing all PFA platforms against RF for reports received August-December 2025 (Supplemental Table S3). Both supplementary analyses used identical search terms, exclusion criteria, and adjudication pipelines as the primary cohort. The two-period design was motivated by the well-characterized Weber effect, wherein regulatory communications inflate adverse event reporting for the highlighted device. The July 2025 cutoff was pre-specified to predate the FDA Safety Communication, isolating the primary analysis from subsequent notoriety-driven reporting inflation. An event-date-based sensitivity analysis was performed for Varipulse to assess whether receipt-date indexing materially affected case inclusion (Supplemental Table S6).

### Note on the Affera Sphere-9 System

The Sphere-9 catheter can deliver both PFA and RF energy. A manual review of event narratives confirmed that 15 of 114 included Affera reports (13.2%) documented pure PFA delivery; the remainder described RF-only, hybrid, or energy-unspecified procedures.

## Results

### Study Cohort

2,862 reports were retrieved from the MAUDE database through July 31, 2025. After excluding 600 reports (Figure 1), 2,262 unique reports remained. This cohort comprised 760 reports from three PFA platforms (Farapulse: 544; Varipulse: 102; Affera: 114) and 1,502 reports from two contemporary RF platforms (QDOT Micro: 967; ThermoCool STSF: 535). The characteristics of the included reports are summarized in Table 1.

**Figure 1.**
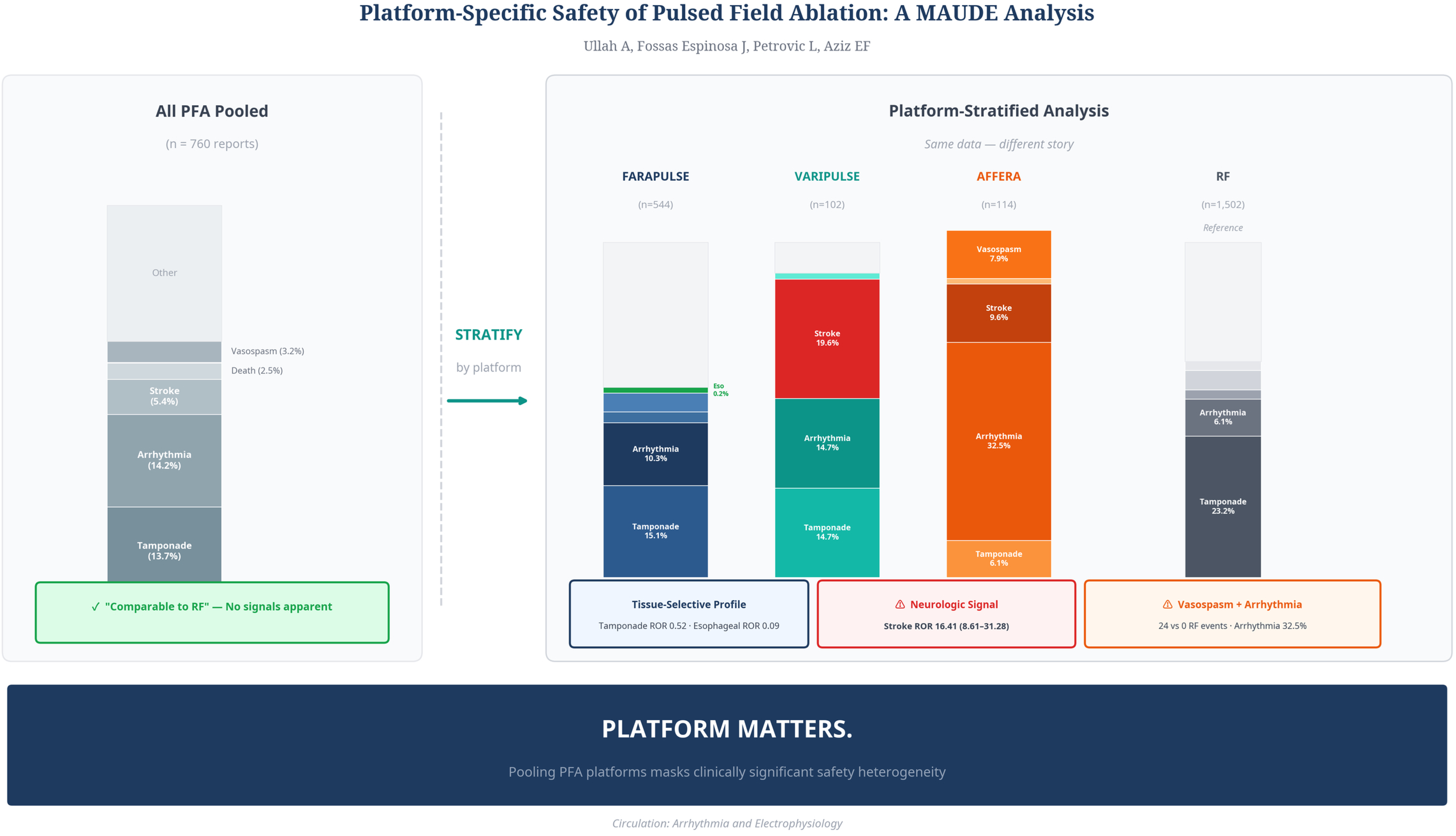
MAUDE Record Identification and Inclusion. Flow diagram yielding 2,262 unique adverse event reports: 760 PFA and 1,502 RF. AF = atrial fibrillation; MAUDE = Manufacturer and User Facility Device Experience; PFA = pulsed-field ablation; RF = radiofrequency.

**Table 1.**
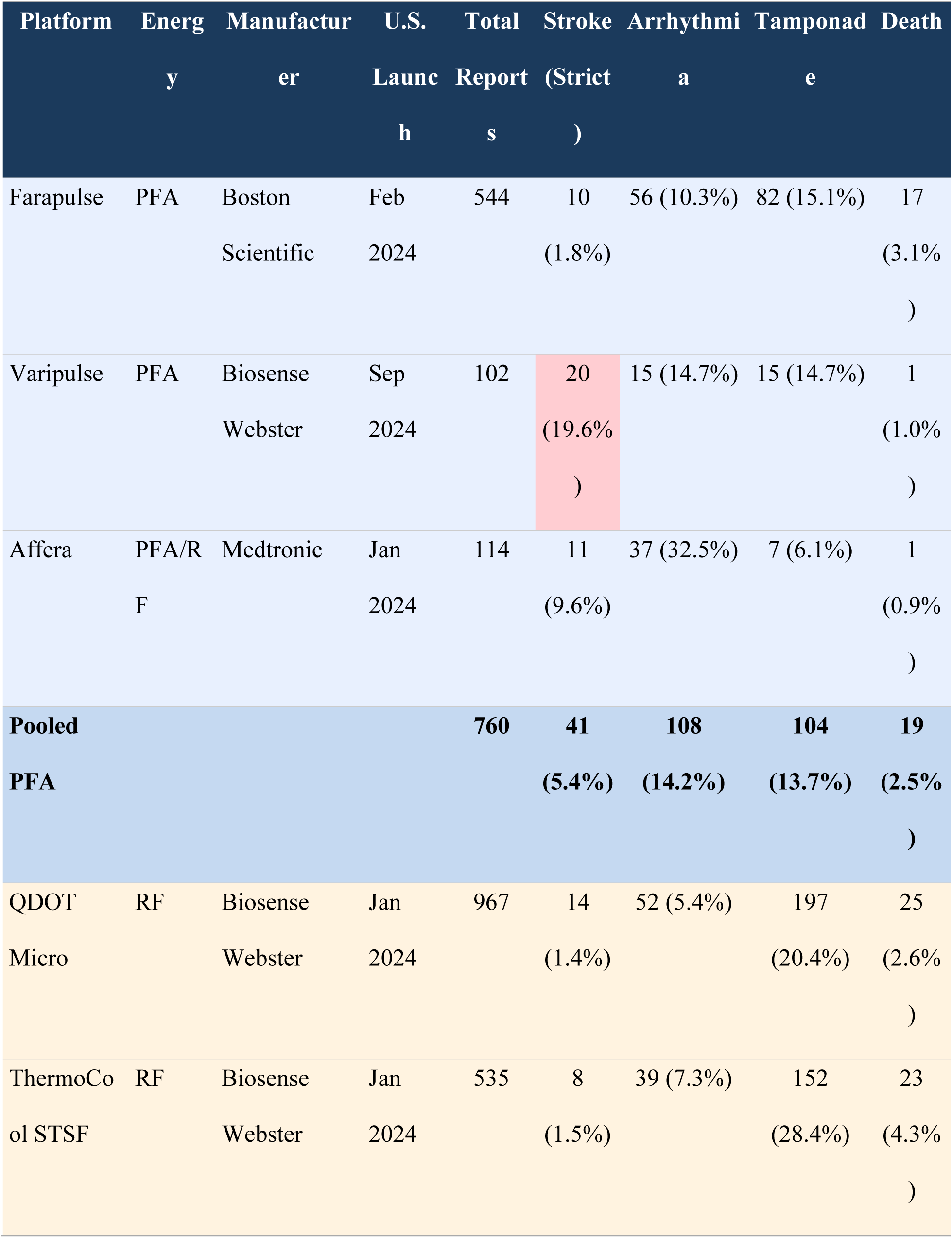
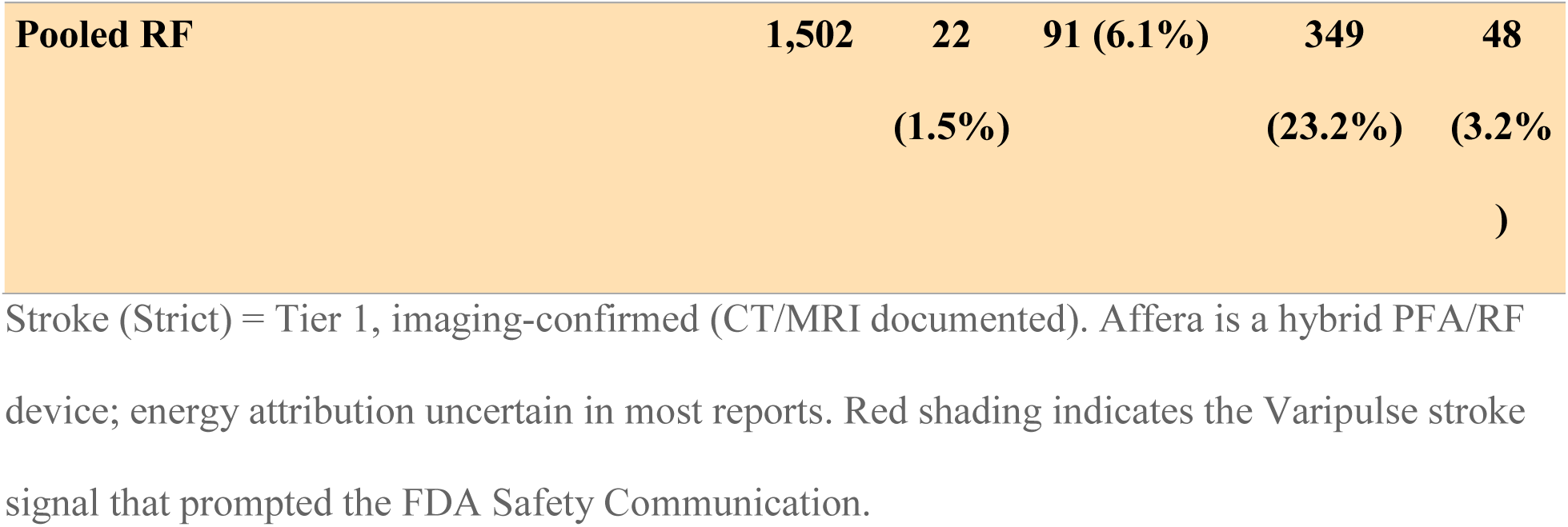
Platform Characteristics and Adverse Event Reporting Proportions (Primary Analysis: January 2024 – July 2025)

### Overall Complication Profile: PFA versus RF

The distribution of adverse events across the nine prespecified complication domains revealed significant differences between the PFA and RF cohorts (Table 2). Disproportionality analysis revealed that, compared to the pooled RF cohort, the pooled PFA cohort had significantly higher reporting odds for imaging-confirmed stroke (ROR 3.84, 95% CI 2.27-6.49) and arrhythmia (ROR 2.57, 95% CI 1.91-3.45). Coronary vasospasm was identified as a PFA-specific signal, with 24 events reported on PFA platforms versus none on RF platforms (p<0.001). Conversely, PFA demonstrated a significantly protective signal, with markedly lower reporting odds for cardiac perforation/tamponade (ROR 0.52, 95% CI 0.41-0.67) and esophageal injury (ROR 0.09, 95% CI 0.01-0.66). No significant differences were observed for death, phrenic nerve injury, or pulmonary vein stenosis after Benjamini-Hochberg correction for multiple comparisons (Figure 3). Narrative review of 67 death reports revealed that cardiac tamponade/perforation was the leading identified mechanism (PFA 4/19 [21.1%] vs. RF 22/48 [45.8%]), while atrioesophageal fistula accounted for 15 RF deaths (31.3%) but zero PFA deaths, consistent with the tissue-selective reporting profile. Insufficient narrative detail precluded cause classification in 10 reports (14.9%), predominantly early Farapulse filings (Supplemental Table S8).

**Table 2.**
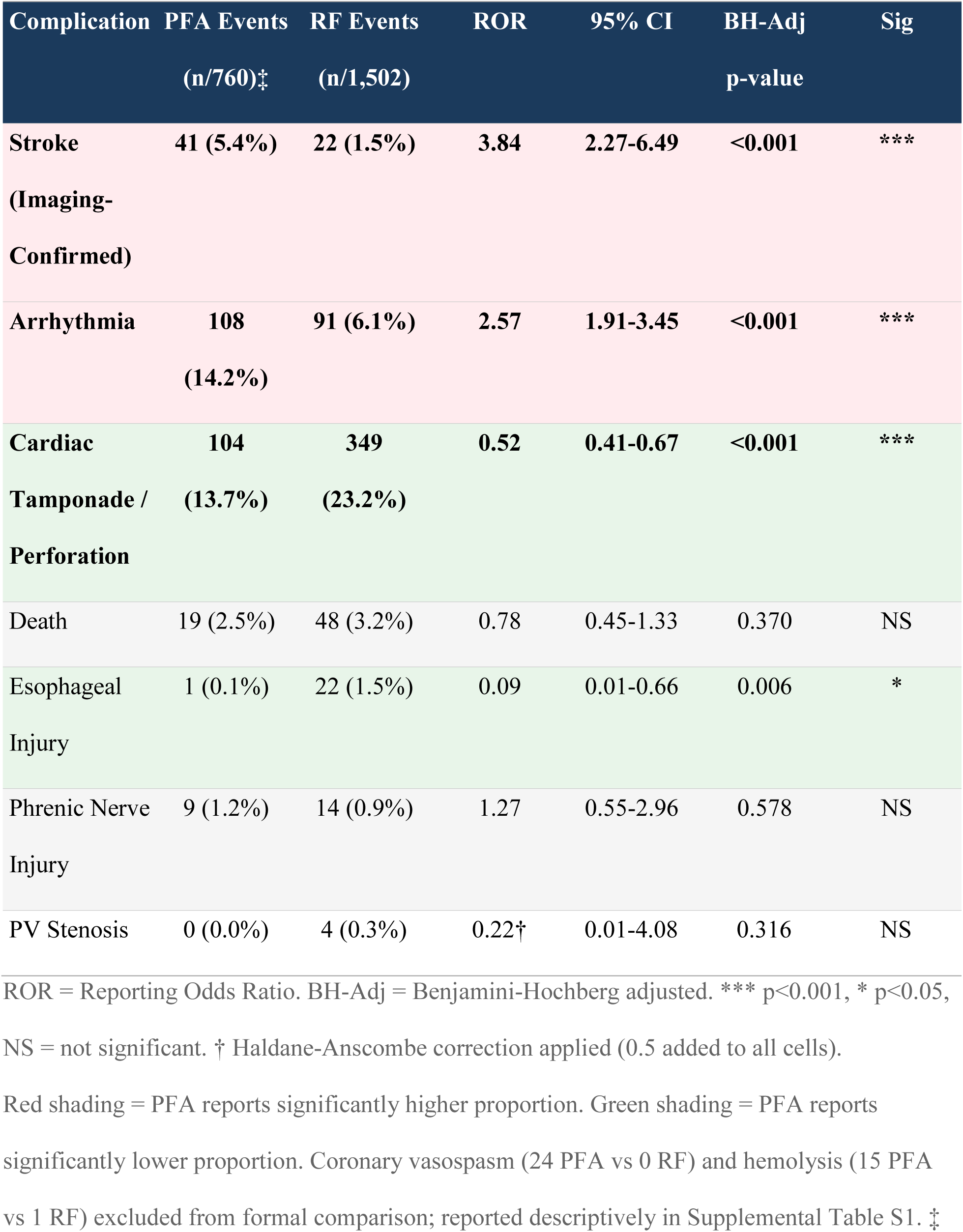
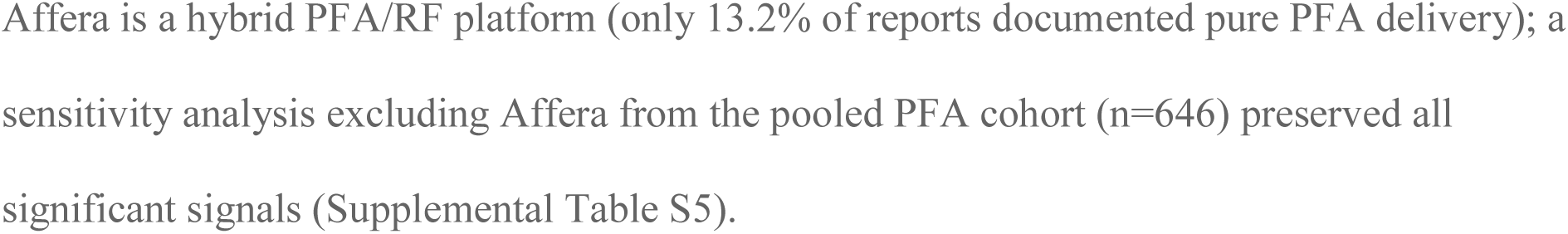
Pooled PFA vs RF Adverse Event Disproportionality Analysis (Reporting Odds Ratios)

| Complication | PFA Events<br>(n/760)<sup>‡</sup> | RF Events<br>(n/1,502) | ROR | 95% CI | BH-Adj<br>p-value | Sig |
| --- | --- | --- | --- | --- | --- | --- |
| Stroke<br>(Imaging-Confirmed) | 41 (5.4%) | 22 (1.5%) | 3.84 | 2.27-6.49 | <0.001 | *** |
| Arrhythmia | 108<br>(14.2%) | 91 (6.1%) | 2.57 | 1.91-3.45 | <0.001 | *** |
| Cardiac<br>Tamponade /<br>Perforation | 104<br>(13.7%) | 349<br>(23.2%) | 0.52 | 0.41-0.67 | <0.001 | *** |
| Death | 19 (2.5%) | 48 (3.2%) | 0.78 | 0.45-1.33 | 0.370 | NS |
| Esophageal<br>Injury | 1 (0.1%) | 22 (1.5%) | 0.09 | 0.01-0.66 | 0.006 | * |
| Phrenic Nerve<br>Injury | 9 (1.2%) | 14 (0.9%) | 1.27 | 0.55-2.96 | 0.578 | NS |
| PV Stenosis | 0 (0.0%) | 4 (0.3%) | 0.22<sup>†</sup> | 0.01-4.08 | 0.316 | NS |

ROR = Reporting Odds Ratio. BH-Adj = Benjamini-Hochberg adjusted. \*\*\* p<0.001, \* p<0.05,
NS = not significant. <sup>†</sup> Haldane-Anscombe correction applied (0.5 added to all cells).
Red shading = PFA reports significantly higher proportion. Green shading = PFA reports
significantly lower proportion. Coronary vasospasm (24 PFA vs 0 RF) and hemolysis (15 PFA
vs 1 RF) excluded from formal comparison; reported descriptively in Supplemental Table S1. <sup>‡</sup>

### Platform-Specific Safety Profiles

Analysis at the individual device level revealed substantial heterogeneity in safety signals, particularly for neurologic events and arrhythmias (Figure 2).

**Figure 2.**
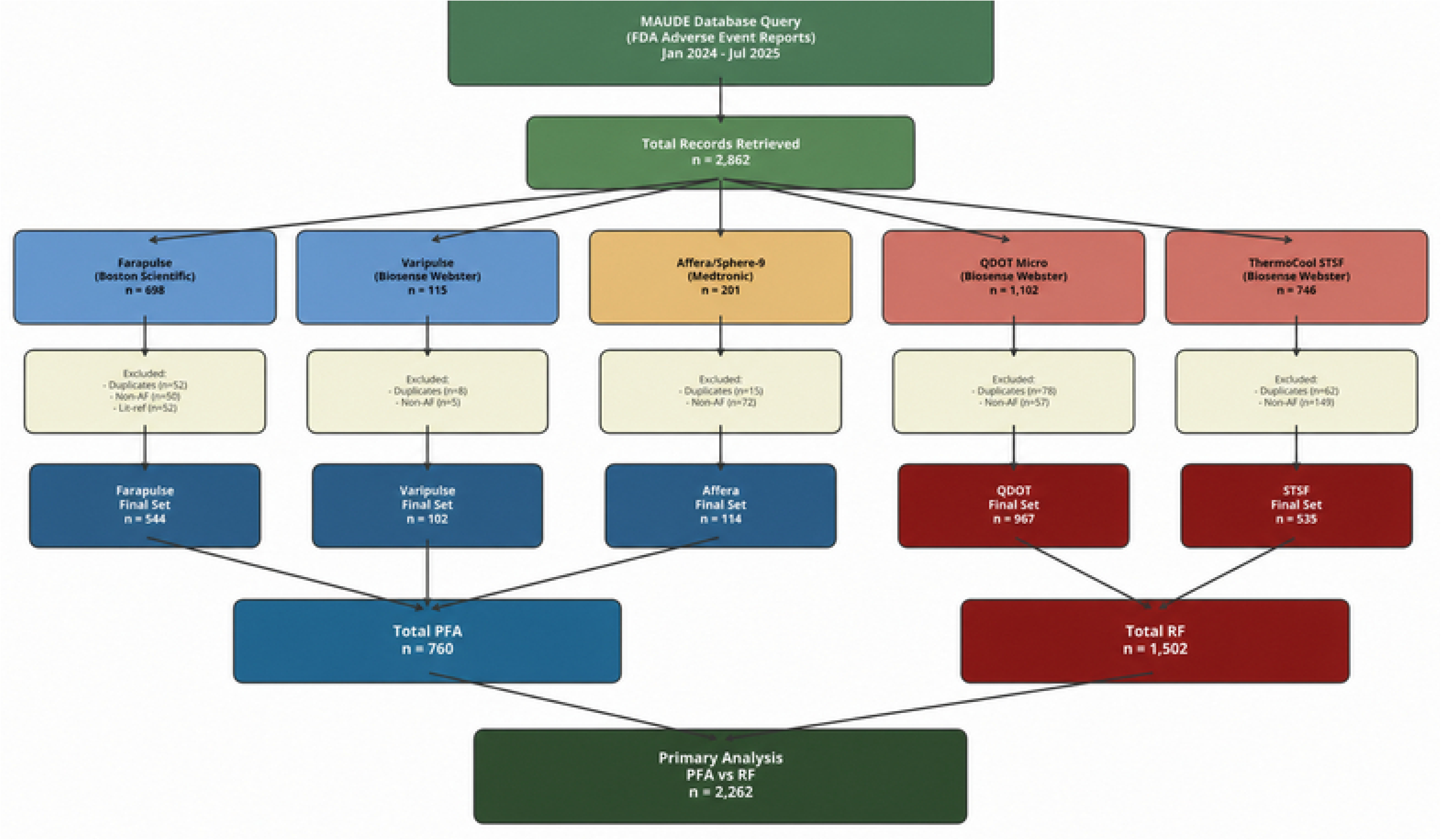
Platform-Specific Complication Profiles. Proportion of MAUDE reports by complication domain and platform, highlighting Varipulse imaging-confirmed stroke reporting and Affera arrhythmia reporting.

**Figure 3.**
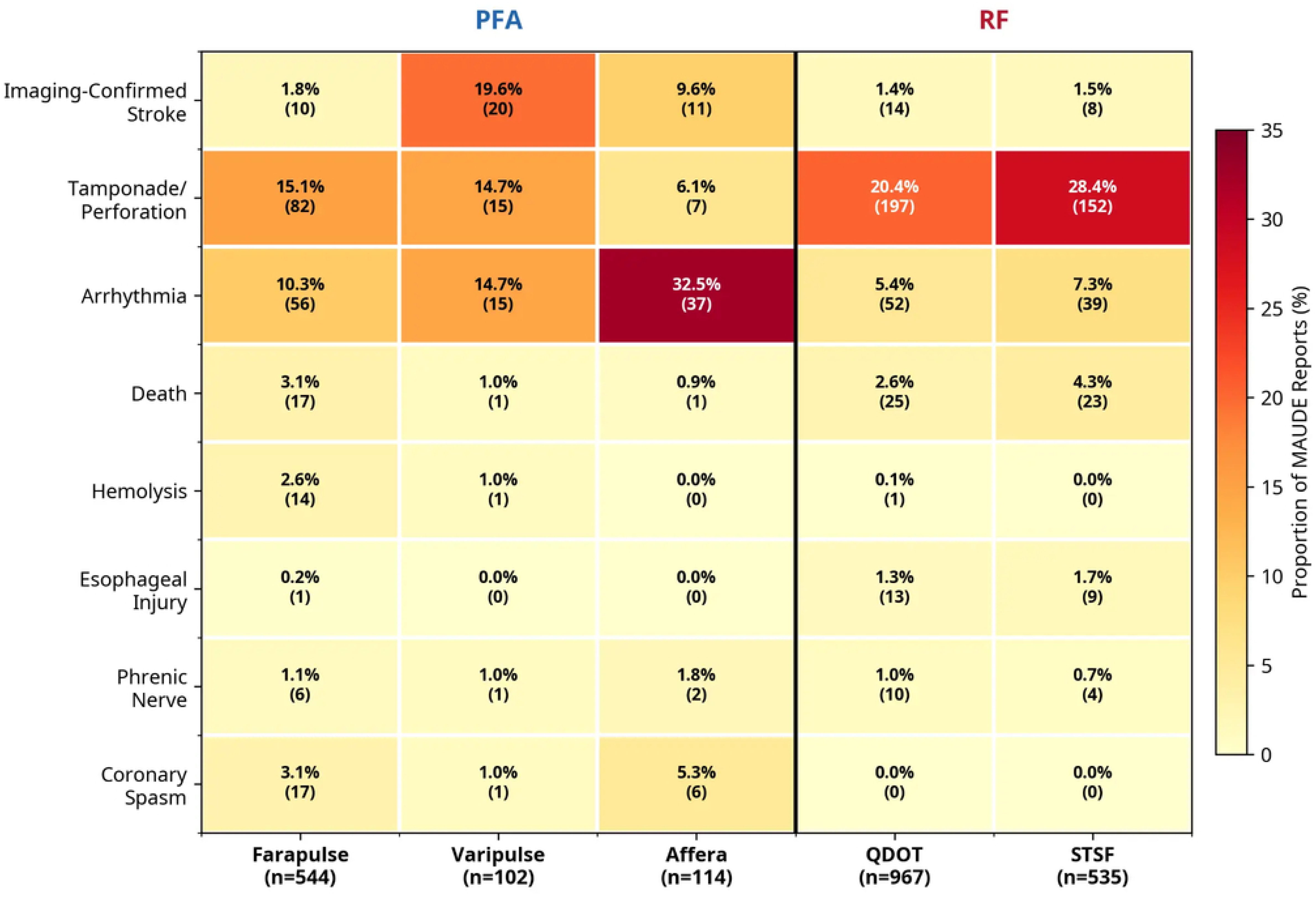
Pooled PFA vs RF Reporting Odds Ratios. RORs and 95% CIs for six mechanism-comparable complication domains. *p<0.05 after BH correction. BH = Benjamini-Hochberg; CI = confidence interval; ROR = reporting odds ratio.

*Stroke (Imaging-Confirmed):* The aggregate PFA stroke signal was driven almost exclusively by the Varipulse platform. Imaging-confirmed strokes were documented in 20 of 102 Varipulse MAUDE reports (19.6% of reports; MAUDE does not capture procedural denominators). This yielded a Reporting Odds Ratio of 16.41 (95% CI 8.61-31.28) when compared with the pooled RF cohort, representing a clear safety signal (Table 3; Figure 4). In contrast, the stroke ROR for the Farapulse platform was non-significant at 1.26 (95% CI 0.59-2.68), with imaging-confirmed events in just 1.8% of reports (10/544). The hybrid-energy Affera platform had a significant ROR of 7.18 (95% CI 3.39-15.22), with events in 9.6% of reports (11/114). Of these 11 imaging-confirmed strokes, only 2 occurred during documented pure-PFA delivery; the remainder involved RF-only, hybrid, or energy-unspecified procedures. Because only 13.2% of Affera reports confirmed pure PFA delivery, this signal is heavily confounded by RF energy and cannot be definitively attributed to PFA. The temporal distribution of Varipulse neurologic events was concentrated in the early commercial period but continued after the manufacturer’s voluntary commercial pause (January 2025) and subsequent IFU revision mandating increased irrigation flow rates (July 2025) (Figure 6).

**Figure 4.**
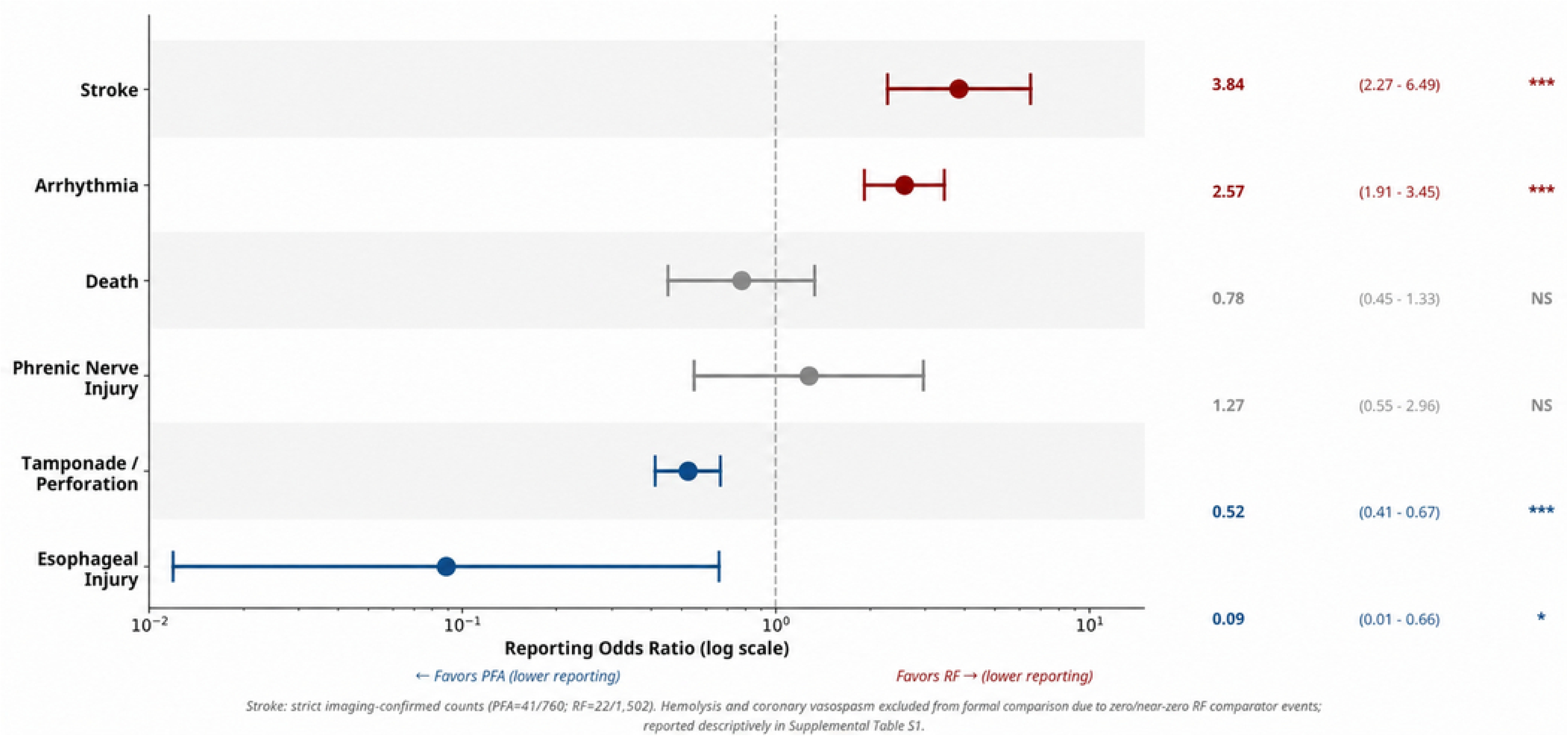
Imaging-Confirmed Stroke ROR by PFA Platform. RORs and 95% CIs for imaging-confirmed stroke versus pooled RF. The neurologic reporting signal is concentrated in Varipulse. CI = confidence interval; ROR = reporting odds ratio.

**Figure 5.**
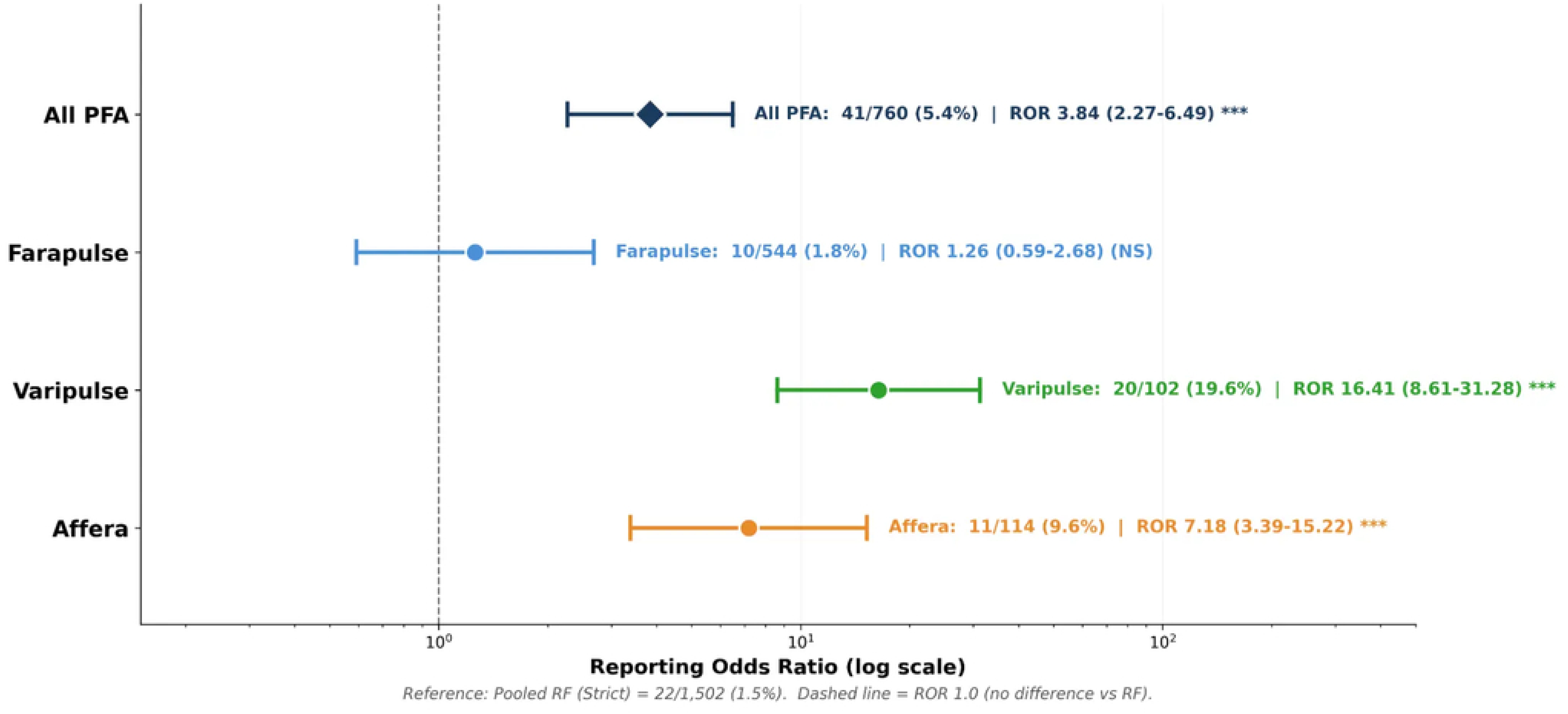
Platform-Specific Safety of PFA Systems. Summary of tissue-selective reporting patterns, platform-specific neurologic reporting, and PFA-specific signals.

**Figure 6.**
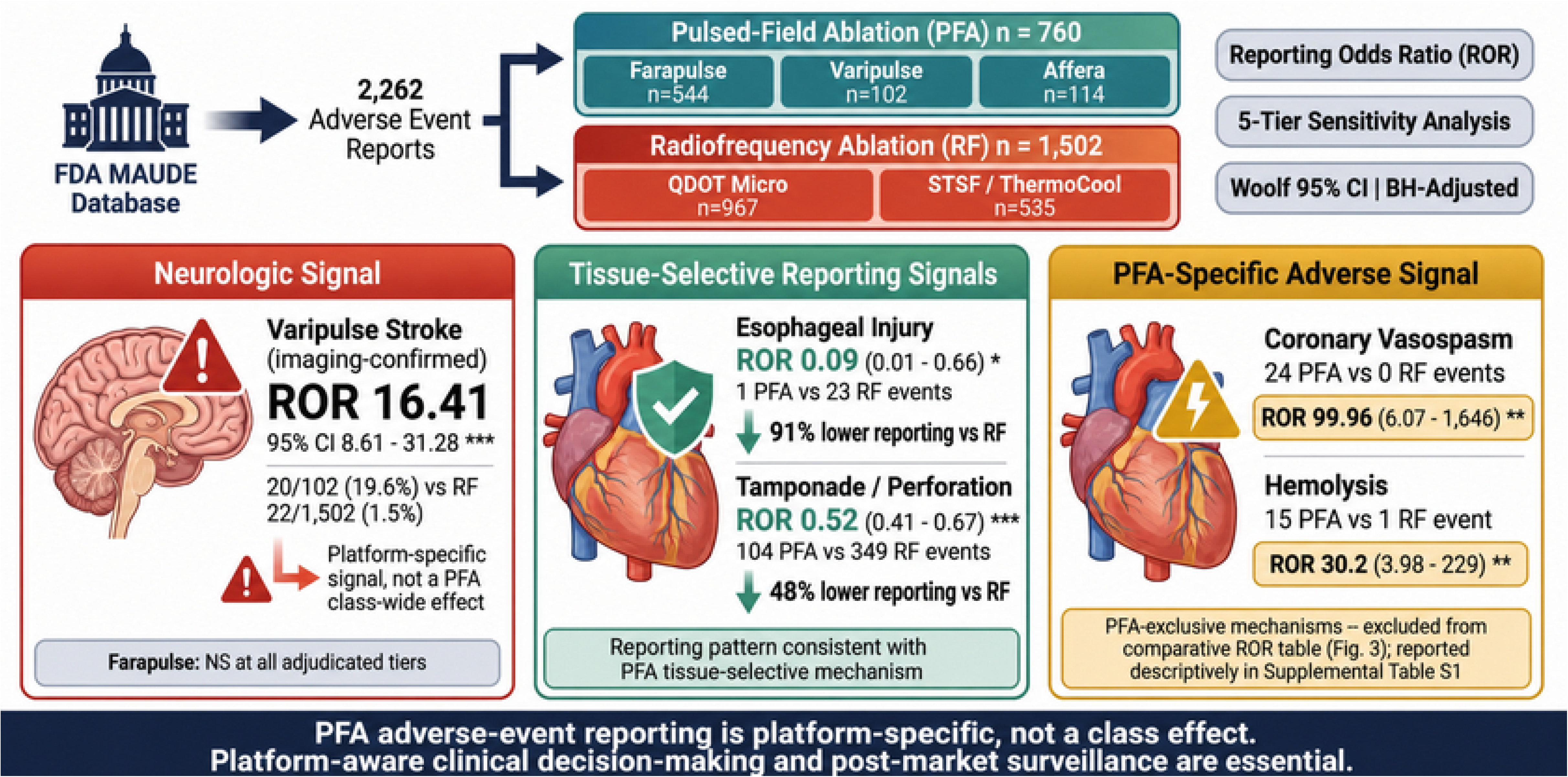
Temporal Distribution of Neurologic Event Reports. Quarterly Varipulse and RF (scaled /10) pre-adjudication neurologic event counts (top) and cumulative events over time (bottom), 2021-2025. The red dashed line indicates the FDA Safety Communication (July 2025); the orange-shaded region denotes the extension period (Q3-Q4 2025). RF counts are scaled (/10) for visual comparability. Q3 2025 spans both study periods. PFA = pulsed-field ablation; RF = radiofrequency.

**Table 3.** Platform-Specific Imaging-Confirmed Stroke Disproportionality vs Pooled RF.

| PFA Platform | N | Stroke T1 Events | Rate | RF Stroke (22/1,502) | ROR | 95% CI | p-value | Sig |
| --- | --- | --- | --- | --- | --- | --- | --- | --- |
| Varipulse | 102 | 20 | 19.6% | 1.5% | 16.41 | 8.61-31.28 | <0.0001 | *** |
| Affera | 114 | 11 | 9.6% | 1.5% | 7.18 | 3.39-15.22 | <0.0001 | *** |
| Farapulse | 544 | 10 | 1.8% | 1.5% | 1.26 | 0.59-2.68 | 0.548 | NS |
| **Pooled PFA** | **760** | **41** | **5.4%** | **1.5%** | **3.84** | **2.27-6.49** | **<0.0001** | **\*\*\*** |

The stroke signal is driven by Varipulse (ROR 16.41) and Affera (ROR 7.18). Farapulse, despite being the most widely reported PFA device (N=544), shows no significant stroke disproportionality (ROR 1.26, NS). Affera is a hybrid PFA/RF device; energy attribution is uncertain in most reports.

Using the pre-adjudication “Original” tier (all MAUDE neurology flags without adjudication), the Varipulse stroke/TIA ROR was 28.43 (95% CI 17.68-45.73), with 52 of 102 total Varipulse reports (51.0%) describing a neurological event. The consistency of the signal across all five adjudication tiers (Supplemental Table S4; Figure 7) indicates that the finding is stable across the definition of neurologic events and is not an artifact of case classification. An event-date sensitivity analysis (97% completeness; median reporting lag 28 days) confirmed that the temporal distribution of Varipulse neurologic events was qualitatively unchanged when indexed to event occurrence date rather than FDA receipt date (Supplemental Table S6). A time-matched comparison of each platform’s first four months of commercial reporting (Farapulse February-May 2024, n=8; Varipulse September-December 2024, n=11) showed that neurologic events were reported in 63.6% of early Varipulse reports versus 0% of early Farapulse reports (ROR 28.33, 95% CI 1.30-617.96, p=0.03; Supplemental Table S7). This signal was robust across all time-matched windows tested (2-6 months), with the ROR strengthening monotonically from 15.00 to 44.73 as the observation window expanded (p=0.009 at 6 months). Month-by-month analysis confirmed that Varipulse neurologic events were present in every month from launch (33-100% of monthly reports), whereas Farapulse reported zero neurologic events during its first 14 months of commercial availability (Supplemental Table S9), indicating a platform-specific signal rather than an early-adoption artifact.

**Figure 7.**
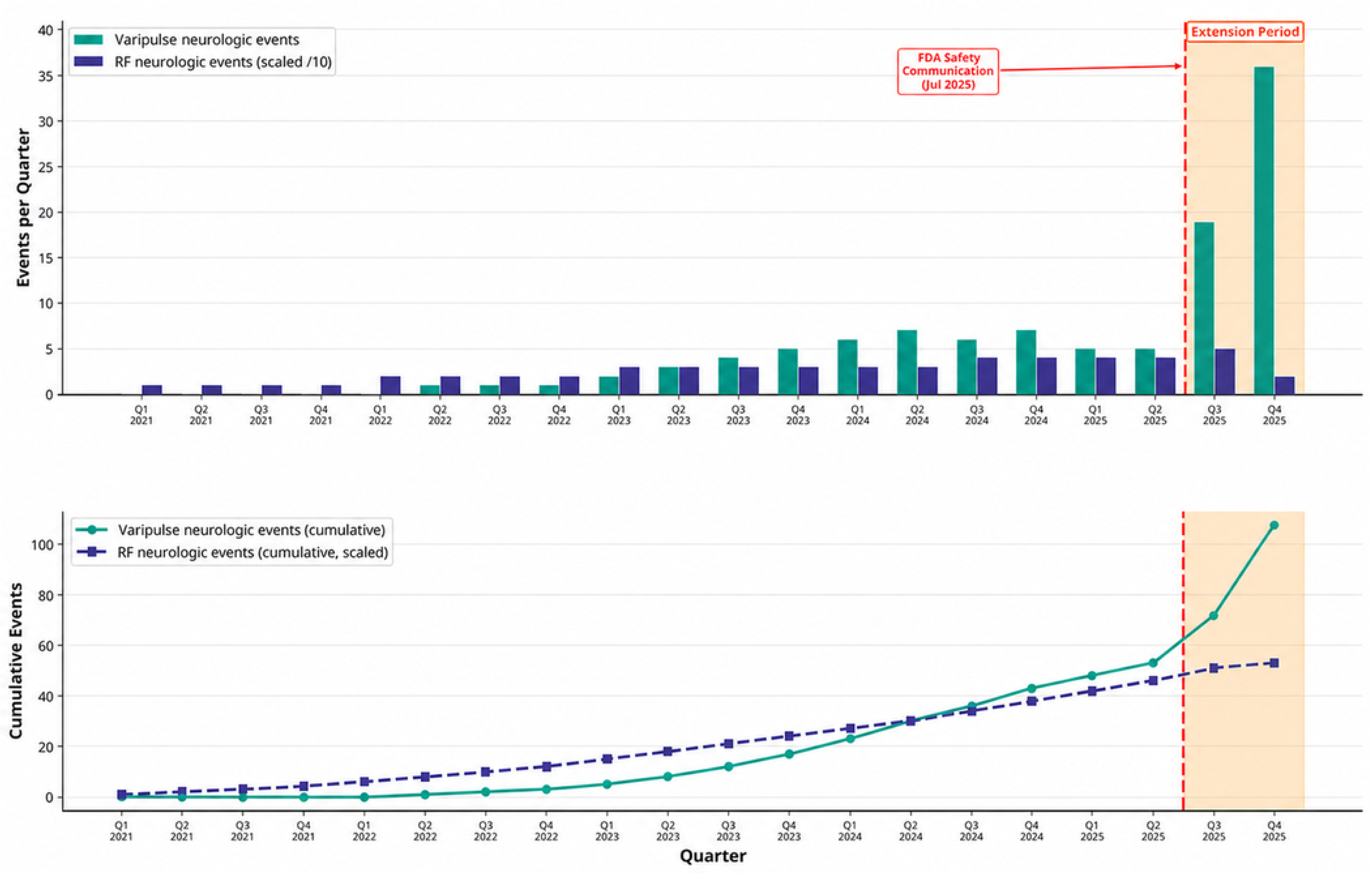
Sensitivity Analysis: Stroke/TIA ROR Across Five Adjudication Tiers. RORs for neurologic events at each of the five adjudication tiers for each PFA platform vs. pooled RF. The Varipulse signal is significant across all tiers. NS = not significant; PFA = pulsed-field ablation; RF = radiofrequency; ROR = reporting odds ratio; TIA = transient ischemic attack.

*Arrhythmia:* The signal for increased arrhythmia reporting in the PFA cohort was disproportionately associated with the Affera Sphere-9 catheter (32.5% of reports, 37/114), compared to 14.7% of Varipulse reports and 10.3% of Farapulse reports. This Affera signal is likely confounded by its hybrid RF capability and VT/VF induction protocols (Figure 2).

*Tissue-Selective Safety:* Tissue-selective protective signals were observed across all PFA platforms. Cardiac perforation/tamponade was reported in a lower proportion of reports for all three PFA platforms (Farapulse 15.1%, Varipulse 14.7%, Affera 6.1%) than for both RF devices (QDOT 20.4%, STSF 28.4%). The near-absence of esophageal injury was particularly notable, with only a single event reported across all 760 PFA reports (0.1%) compared to 22 events in the RF cohort (1.5%).

*PFA-Specific Complications:* Coronary vasospasm was reported exclusively in PFA cases, with 17 events in Farapulse, 6 in Affera, and 1 in Varipulse. Hemolysis was also predominantly PFA-specific, with 14 of the 15 PFA events occurring in Farapulse reports. Because only a single hemolysis event was reported in the pooled RF cohort, formal disproportionality analysis yields an exceptionally wide confidence interval; therefore, this finding is best interpreted descriptively as a PFA-specific signal.

### Extension Period Validation

In the pre-specified extension period analysis (August-December 2025), a total of 1,414 PFA and 402 RF reports were evaluated (Supplemental Table S3). This secondary cohort provided a natural experiment to test the stability of the primary findings following the July 2025 FDA Safety Communication. The pooled PFA imaging-confirmed stroke signal attenuated from a highly significant ROR of 3.84 in the primary analysis to non-significance in the extension period (ROR 1.60, 95% CI 0.89-2.85, p=0.11). This attenuation is compatible with notoriety bias, wherein the regulatory alert stimulated disproportionate reporting of non-neurological adverse events across all PFA platforms, thereby diluting the relative stroke signal.

Despite this shift in reporting behavior, the core tissue-selective reporting patterns of PFA were sustained. PFA maintained significantly lower reporting odds for cardiac tamponade (ROR 0.48, 95% CI 0.37-0.62, p<0.0001) and death (ROR 0.44, 95% CI 0.30-0.65, p<0.0001) in the extension cohort. Furthermore, the PFA-specific complications identified in the primary analysis persisted: coronary vasospasm (63 vs. 1 event; ROR 18.70, 95% CI 2.59-135.25, p=0.004) and hemolysis (25 vs. 0 events; p=0.003) remained highly specific to pulsed-field energy. The reproducibility of these specific signals across both study periods reinforces that they support classification as mechanism-related pharmacovigilance signals rather than transient artifacts of stimulated reporting.

### Composite Serious Adverse Events

In the pre-specified primary composite safety endpoint (stroke/TIA, pericardial tamponade or perforation, death, phrenic nerve injury, and esophageal injury), pulsed-field and radiofrequency platforms demonstrated comparable reporting proportions (PFA 229/760 [30.1%] vs. RF 486/1,502 [32.4%]; ROR 0.90, 95% CI 0.75-1.09, p=0.280). Mechanism-specific PFA complications (hemolysis, coronary vasospasm, pulmonary vein stenosis) were reported descriptively and excluded from the composite because no stable RF comparator exists. The absence of a composite difference reflects opposing signals: the significantly higher stroke reporting for Varipulse was offset by the significantly lower tamponade and esophageal injury reporting across the PFA class.

## Discussion

This analysis of 2,262 manually adjudicated MAUDE adverse event reports is, to our knowledge, the first to systematically compare the adverse-event reporting profiles of all three FDA-approved PFA platforms against contemporary RF ablation catheters using a rigorous, multi-auditor adjudication process for neurologic events. The study yields three principal findings. First, PFA was associated with a tissue-selective reporting profile, with lower reporting of tamponade and esophageal injury compared with RF. Second, PFA adverse-event reporting demonstrates substantial platform heterogeneity rather than a uniform class-wide reporting profile; the neurologic signal for PFA is driven almost entirely by the Varipulse platform and is not observed with Farapulse. Third, PFA reporting reveals distinct, non-thermal safety signals, including coronary vasospasm and hemolysis, that are not observed with RF ablation.

### PFA Tissue Selectivity: Post-Market Reporting Evidence

The primary analysis is consistent with the foundational premise of pulsed-field ablation, superior tissue selectivity, established in preclinical studies. Only a single PFA-associated esophageal event was reported in the primary cohort (a Farapulse case that also involved coincident RF energy delivery), compared to 22 events in the RF cohort. This aligns with preclinical data showing a higher electroporation threshold for esophageal wall than myocardium [2, 4, 18].

This lower tamponade reporting signal is plausibly attributable, at least in part, to the non-thermal nature of PFA, the elimination of steam pops (which can cause acute perforation during RF delivery), and shorter dwell times. The consistency of this protective signal across both study periods and across all PFA platforms is consistent with the hypothesis that reduced mechanical trauma is an inherent property of pulsed-field energy delivery rather than an artifact of differential reporting.

### The Varipulse Neurological Signal: A Case Study in Pharmacovigilance

The Varipulse stroke/TIA ROR of 28.43 (Original tier) and imaging-confirmed stroke ROR of 16.41 (Strict tier) versus RF represent strong pharmacovigilance signals. That 51.0% of all Varipulse reports describe a neurological event (Original tier), compared with 3.5% of RF reports, is difficult to attribute to stimulated reporting alone, although this cannot be fully separated from notoriety bias in MAUDE. This signal is corroborated by a prospective MRI study demonstrating ischemic cerebral lesions in 66.7% of patients undergoing VLCC ablation at original protocol [9], and the manufacturer’s commercial complaint database documenting a 3.0% pre-pause neurovascular event frequency in the manufacturer commercial experience [13].

The underlying mechanism has been characterized in preclinical and clinical studies: at the original 4 mL/min irrigation rate, VLCC electrodes undergo Joule heating during high-voltage pulse delivery, reaching up to 86 degrees C [14]. This superheating produces embolic hazards including large gas microbubbles, hemolysis, and char formation. Revision of the IFU to require 30 mL/min irrigation markedly suppresses heat stacking and large bubble formation [10, 11, 12, 15], and was followed by a 10-fold reduction in neurovascular event frequency [13]. Our MAUDE dataset primarily captures the pre-pause commercial era.

These neurological events represent a discrete, potentially addressable safety issue tied to a specific procedural protocol that has since been modified. To evaluate signal stability, we performed a supplementary analysis of 202 Varipulse reports received after the July 2025 IFU revision (Supplemental Table S10). Applying identical exclusion and adjudication criteria, the imaging-confirmed stroke proportion remained elevated at 14.6% (26/178 cleaned reports), compared to 1.2% for contemporary RF (8/646 cleaned reports). While this represents a modest reduction from the pre-IFU reporting proportion (19.6%), it remains elevated relative to RF. Interpretation of this temporal trend is limited by potential stimulated reporting bias (notoriety bias) following the manufacturer’s safety communication, which may affect both stroke and non-stroke reporting. A multicenter registry of 1,014 VLCC cases under the revised protocol reported no strokes or coronary spasm [25]. Ongoing surveillance is required to confirm whether this favorable profile is representative of the revised IFU era.

### The Weber Effect and Notoriety Bias

The temporal distribution of adverse event reporting demonstrates the Weber effect (notoriety bias). Following the July 2025 FDA Safety Communication, Varipulse imaging-confirmed stroke reporting proportion rose from 19.6% (primary period) to 31.1% (extension period). This divergence is illustrated in Figure 6. Conversely, while Farapulse stroke reporting increased slightly in absolute terms, it fell below the RF comparator proportion (2.3% vs. 3.5%) during the extension, suggesting stimulated reporting was specific to the highlighted platform. Furthermore, the strong protective PFA esophageal injury signal (ROR 0.09) lost significance when extension data were pooled (combined ROR 1.08), as heightened scrutiny likely prompted reporting of any adverse outcome. Our two-period design isolates the baseline disproportionality signal from subsequent notoriety bias, suggesting elevated stroke reporting is a platform-specific phenomenon. This platform specificity is further supported by month-by-month analysis: Farapulse reported zero neurologic events during its first 14 months of commercial availability (February 2024-March 2025; 47 reports), whereas Varipulse reported neurologic events in every month from launch, with monthly proportions ranging from 33% to 100% (Supplemental Table S9). If the Varipulse signal were an early-adoption artifact, a comparable pattern should have appeared during the Farapulse early-adoption period; it did not.

The attenuation of the pooled stroke ROR from 3.84 (primary) to 1.60 (extension, non-significant) likely reflects both genuine risk reduction from the concurrent IFU revision (increasing irrigation from 4 to 30 mL/min) and notoriety bias inflating the reporting denominator; these two effects cannot be fully disentangled in this dataset. This attenuation illustrates that regulatory alerts can rapidly alter spontaneous-reporting database composition, making temporally unstratified post-alert MAUDE analyses difficult to interpret.

### Coronary Vasospasm: An Emerging PFA-Class Signal

The identification of 24 coronary vasospasm events exclusively in the PFA group (with zero RF events) represents a notable pharmacovigilance signal not captured in controlled trials. The likely mechanism is direct high-voltage field stimulation of coronary smooth muscle, analogous to skeletal muscle contraction during PFA delivery. Preclinical studies confirm that electric field strengths sufficient for electroporation can induce transient coronary vasospasm in adjacent vessels [14, 15]. Delayed vasospasm up to 45 minutes post-ablation has been reported [26].

While generally transient and responsive to intracoronary nitroglycerin, coronary vasospasm can cause hemodynamic compromise, ST elevation, and procedural termination. The persistence of this signal in the extension period (63 events, ROR 18.70) supports its classification as a mechanism-related pharmacovigilance signal. Centers performing high-volume PFA may consider including coronary vasospasm in periprocedural monitoring protocols, with access to intracoronary nitrates and coronary angiography when clinically indicated.

### Clinical Implications

The informed consent process for atrial fibrillation ablation may benefit from becoming platform-specific rather than treating all PFA systems as equivalent. Patients may benefit from counseling on the specific catheter’s reported risk profile. Varipulse operators should follow the revised Instructions for Use regarding sheath management and irrigation protocols, and should maintain a low threshold for periprocedural neuroimaging if any neurological deficits are suspected. Electrophysiology laboratories transitioning to high-volume PFA programs should be prepared to recognize and manage coronary vasospasm, with intracoronary nitroglycerin readily available.

Finally, while these reporting signals require vigilance, the substantially lower reporting of esophageal injury and cardiac tamponade support the continued adoption of PFA technology. The MANIFEST-17K registry [19] reports a Farapulse stroke incidence of approximately 0.4% (from a prospective registry with procedural denominators [19]), versus a reporting proportion of 1.8% in our MAUDE cohort, underscoring that disproportionality signals reflect relative reporting patterns, not absolute incidence. The value of this analysis lies in identifying platform-differentiated signals requiring targeted clinical attention.

### Limitations

MAUDE is a passive surveillance database subject to underreporting and stimulated reporting bias. It lacks procedural denominators, precluding absolute incidence rates; ROR measures relative disproportionality, not absolute risk. Importantly, disproportionality analysis is a signal-generation method designed to identify reporting imbalances warranting further investigation; elevated RORs do not establish causality, and the absence of reliable denominators means that reporting proportions cannot be equated with clinical event rates. All findings should therefore be interpreted as hypothesis-generating and require confirmation through prospective registries or controlled studies with known procedural volumes. Reports lack consistent demographic data, precluding sex-stratified analysis.

As Dhruva and Shivkumar have noted [22], MAUDE is positioned to detect rare signals before they accumulate in registry or trial data.

The Affera hybrid energy platform presents a specific limitation: because only 13.2% of reports explicitly documented pure PFA delivery, attribution of complications specifically to PFA energy cannot be definitively confirmed for the majority of Affera reports. A sensitivity analysis excluding all Affera reports from the pooled PFA cohort (n=646 vs. RF n=1,502) preserved the direction and statistical significance of all primary findings (Supplemental Table S5), confirming that the core disproportionality signals are not driven by the hybrid platform.

All three adjudicators are from a single institution, which may introduce systematic bias in event classification; however, the high inter-auditor agreement (Fleiss’ kappa = 0.92) and the pre-specified five-tier framework mitigate this concern. The Varipulse dataset spans both the high-signal pre-pause era and the early post-pause IFU-revised era, meaning the aggregate ROR may not reflect the current reporting profile under the revised protocol. Furthermore, MAUDE reports frequently lack granular procedural details, such as ACT levels, anticoagulation protocols, sheath management, and operator experience. The RF comparators (QDOT Micro and ThermoCool STSF) were selected as the most contemporary, high-volume irrigated-tip RF platforms from the same manufacturer (Biosense Webster) to control for reporting culture, as manufacturer-specific field safety teams, complaint-processing workflows, and MDR submission practices can influence reporting volume and granularity independent of actual event frequency. Inclusion of RF platforms from different manufacturers would introduce reporting-infrastructure confounding that cannot be adjusted for in MAUDE analyses. Other RF catheters (e.g., TactiFlex) were not included. These unmeasured confounders, along with institutional learning curve effects during early PFA adoption, cannot be adjusted for in this dataset.

## Clinical Perspective

### What Is Known

- Three pulsed-field ablation platforms are FDA-approved for atrial fibrillation, but whether their post-market safety profiles differ from each other and from radiofrequency ablation has not been systematically evaluated using device-specific pharmacovigilance data.
- The FDA MAUDE database enables signal detection for rare adverse events not captured in pivotal trials, but prior PFA analyses evaluated these platforms as a single class without platform-level stratification.

### What the Study Adds

- Pulsed-field ablation demonstrates a tissue-selective reporting profile with substantially lower esophageal injury and cardiac tamponade reporting compared with radiofrequency ablation, consistent across all three platforms.
- The Varipulse neurologic reporting signal (reporting odds ratio 16.41) is platform-specific and not observed with Farapulse (reporting odds ratio 1.26), indicating that pulsed-field ablation safety data should not be pooled across devices.
- Coronary vasospasm is a pulsed-field ablation class-specific signal (24 vs. 0 radiofrequency events) requiring periprocedural awareness.

## Conclusions

PFA adverse-event reporting was heterogeneous across approved platforms, supporting platform-level interpretation of post-market safety signals. PFA displays a tissue-selective reporting profile relative to RF, with near-absence of reported esophageal injury and lower tamponade reporting. The Varipulse catheter exhibited a marked neurological event signal during its initial commercial introduction, consistent with a Joule-heating pathway characterized in prior mechanistic studies and subsequently addressed through IFU modifications. Coronary vasospasm constitutes an emerging PFA class signal. These findings support a platform-aware approach to PFA adoption and underscore the ongoing necessity of rigorous post-market surveillance. Future regulatory submissions, society guidelines [23], and clinical trial designs may benefit from platform-level stratification by device architecture and pulse parameters.

## Acknowledgments

None.

## Author Contributions

E.F. Aziz: Conceptualization, Supervision, Writing – Review & Editing. A. Ullah: Conceptualization, Methodology, Data Curation, Formal Analysis, Investigation, Visualization, Writing – Original Draft. J. Fossas Espinosa: Investigation, Data Curation, Writing – Review & Editing. L. Petrovic: Investigation, Data Curation, Writing – Review & Editing.

## Data Availability Statement

All data used in this study are publicly available through the FDA MAUDE database (https://www.accessdata.fda.gov/scripts/cdrh/cfdocs/cfmaude/search.cfm).

## Nonstandard Abbreviations and Acronyms

BH: Benjamini-Hochberg
IFU: instructions for use
MAUDE: Manufacturer and User Facility Device Experience
PFA: pulsed-field ablation
RF: radiofrequency
ROR: reporting odds ratio
SAE: serious adverse event

## Tables

Pulsed Field vs Radiofrequency Ablation Safety: A MAUDE Database Disproportionality Analysis

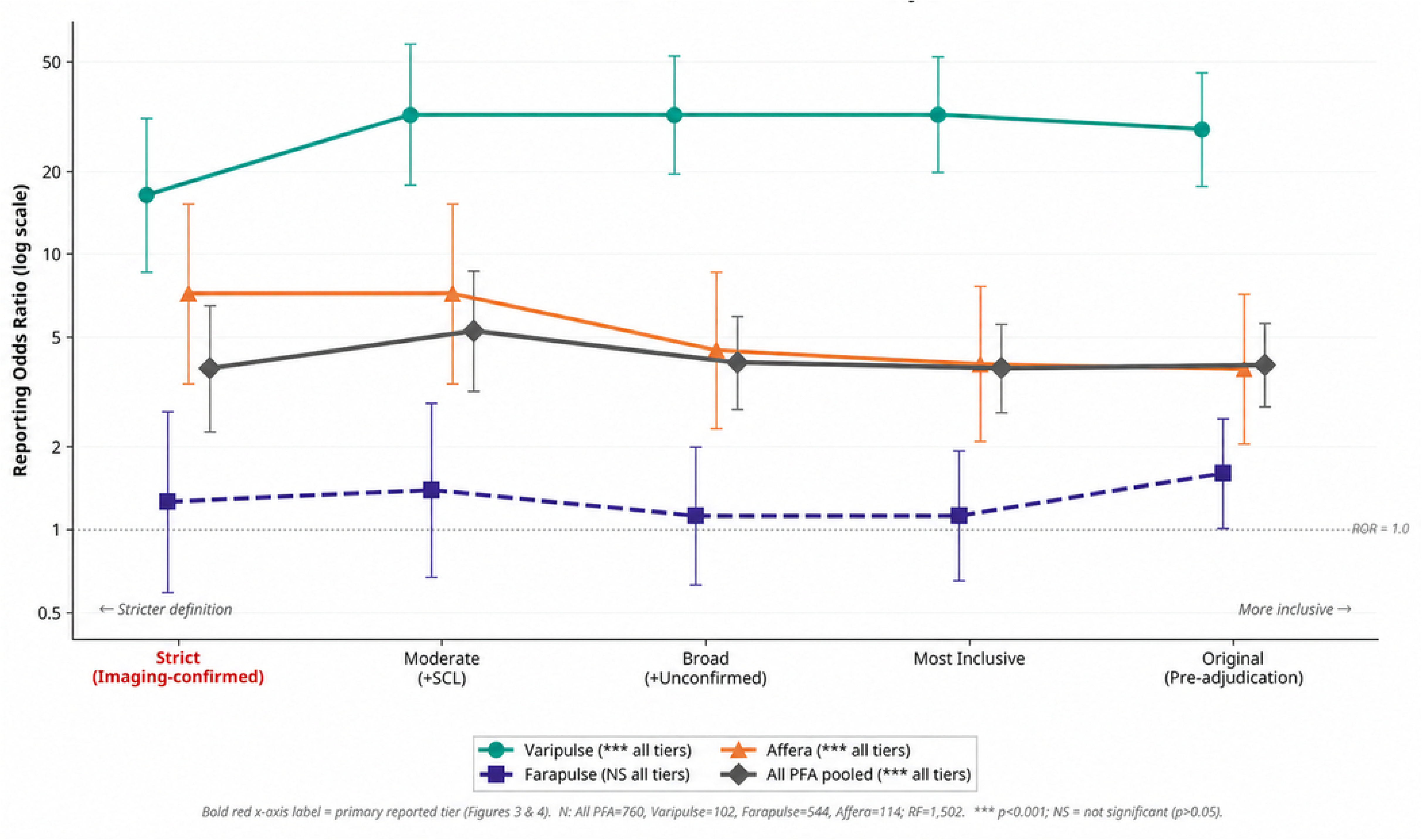

## References

1. von Elm E, Altman DG, Egger M, et al. The Strengthening the Reporting of Observational Studies in Epidemiology (STROBE) statement: guidelines for reporting observational studies. Ann Intern Med. 2007;147(8):573–577.

2. Koruth JS, Kuroki K, Iwasawa J, et al. Pulsed Field Ablation Versus Radiofrequency Ablation: Esophageal Injury in a Novel Porcine Model. Circ Arrhythm Electrophysiol. 2020;13(3):e008303.

3. Reddy VY, Neuzil P, Koruth JS, et al. Pulsed Field Ablation for Pulmonary Vein Isolation in Atrial Fibrillation. J Am Coll Cardiol. 2019;74(3):315–326.

4. Turagam MK, Neuzil P, Schmidt B, et al. Safety and Effectiveness of a Single-Shot Pulsed Field Ablation Catheter for the Treatment of Atrial Fibrillation: A Prospective, Multicenter, Real-World Registry (MANIFEST-PF). Circulation. 2023;148(1):35–46.

5. U.S. Food and Drug Administration. Manufacturer and User Facility Device Experience (MAUDE) Database. Available at: https://www.accessdata.fda.gov/scripts/cdrh/cfdocs/cfmaude/search.cfm. Accessed August 2025.

6. Cho MS, Lee SR, Black-Maier E, et al. Complications Associated with Pulsed Field Ablation vs Radiofrequency Catheter Ablation of Atrial Fibrillation. Heart Rhythm. 2025;22(9):2194–2200. doi:10.1016/j.hrthm.2024.10.063

7. Calkins H, Hindricks G, Cappato R, et al. 2017 HRS/EHRA/ECAS/APHRS/SOLAECE expert consensus statement on catheter and surgical ablation of atrial fibrillation. Heart Rhythm. 2017;14(10):e275–e444.

8. van Puijenbroek EP, Bate A, Leufkens HGM, et al. A comparison of measures of disproportionality for signal detection in spontaneous reporting systems for adverse drug reactions. Pharmacoepidemiol Drug Saf. 2002;11(1):3–10.

9. Laskov V, Hozman M, Malikova H, et al. Cerebrovascular Ischemic Lesions After Pulsed Field Ablation for Atrial Fibrillation Using Variable-Loop Ablation Catheter. J Am Coll Cardiol EP. 2026;12(4):814–823. doi:10.1016/j.jacep.2025.12.018

10. Zhou D, Li M, Song Z, et al. Impact of saline irrigation on haemolysis, silent cerebral lesion incidence, thermal dynamics, and bubble formation in pulsed field ablation with a variable-loop circular catheter. EP Europace. 2026;28(2):euag005. doi:10.1093/europace/euag005

11. Nakahara S, Aoki H, Sato H, et al. Irrigation Flow Rate and Silent Cerebral Events During Pulsed Field Ablation With a Variable-Loop Catheter. J Am Coll Cardiol EP. 2026;12(1). doi:10.1016/j.jacep.2025.09.007

12. Yamashita K, Kikuchi Y, Yoshiyama K, et al. Evaluation of the Impact of Irrigation Flow Rate on Clinical Outcomes During Pulsed Field Ablation. J Cardiovasc Electrophysiol. 2026;37(1):181–185. doi:10.1111/jce.70194

13. Mansour M, Michaud G, Di Biase L, et al. Impact of Lesion Delivery and Irrigation Rates on Neurovascular Events With a Variable Loop Circular Catheter for Pulsed Field Ablation of Atrial Fibrillation. J Am Coll Cardiol EP. 2026. doi:10.1016/j.jacep.2026.01.025

14. Sauer WH, Campos-Villarreal D, Steiger NA. Irrigation of Pulsed Field Ablation Electrodes Mitigates Joule Heating and the Heat Stacking Phenomena. J Am Coll Cardiol EP. 2025;11(8). doi:10.1016/j.jacep.2025.04.018

15. Sauer WH, Coll M, Campos-Villarreal D, Steiger NA. Effect of Electrode Cooling with High Irrigation Rate on Pulsed Field Ablation Lesion Characteristics. Heart Rhythm. 2025 Dec 26. doi:10.1016/j.hrthm.2025.12.034

16. Hoffman KB, Dimbil M, Erdman CB, et al. The Weber Effect and the United States Food and Drug Administration’s Adverse Event Reporting System (FAERS): Analysis of Sixty-Two Drugs Approved from 2006 to 2010. Drug Saf. 2014;37(4):283–294.

17. Pariente A, Gregoire F, Fourrier-Reglat A, et al. Impact of Safety Alerts on Measures of Disproportionality in Spontaneous Reporting Databases: The Notoriety Bias. Drug Saf. 2007;30(10):891–898.

18. Schmidt B, Bordignon S, Tohoku S, et al. EUropean real-world outcomes with Pulsed field ablatiOn in patients with symptomatic atRIal fibrillation: lessons from the multi-centre EU-PORIA registry. Europace. 2023;25(7):euad185. doi:10.1093/europace/euad185

19. Ekanem E, Reddy VY, Schmidt B, et al. Safety of Pulsed Field Ablation in More Than 17,000 Patients with Atrial Fibrillation in the MANIFEST-17K Study. Nat Med. 2024;30(7):2020–2029.

20. Tilz RR, Pürerfellner H, Kuck KH, et al. Underreporting of Complications Following AF Ablation: Comparison of the Manufacturer and User Facility Device Experience FDA Database and a Voluntary Invitation-Based Registry: The POTTER-AF 3 Study. Heart Rhythm. 2025;22(6):1472–1479. doi:10.1016/j.hrthm.2024.09.060

21. Reddy VY, Gerstenfeld EP, Natale A, et al. Pulsed Field or Conventional Thermal Ablation for Paroxysmal Atrial Fibrillation. N Engl J Med. 2023;389(18):1660–1671.

22. Dhruva SS, Kadakia KT, Shivkumar K. Postmarket Data-Real World Safety Evaluation of Pulsed Field Ablation-and Beyond. J Am Coll Cardiol. 2026;87(2):197–199. doi:10.1016/j.jacc.2025.10.061

23. Verma A, Hocini M, Andrade J, et al. 2026 HRS/EHRA Scientific Statement on Pulsed Field Ablation for Cardiac Arrhythmias. Heart Rhythm. 2026. doi:10.1016/j.hrthm.2026.02.006 [Epub ahead of print]

24. Kühne M, Badertscher P, Andrade JG, et al. Pulsed Field Ablation for the Interventional Treatment of Atrial Fibrillation: A Scientific Statement of the EHRA, HRS, APHRS, LAHRS and CHRS. Europace. 2026;euag080. doi:10.1093/europace/euag080

25. Porterfield C, Dominic P, Khaykin Y, et al. Variable loop circular catheter pulsed field ablation in real-world practice: Low complication rates across patient and procedural characteristics in the REAL AF registry. Heart Rhythm. 2026. doi:10.1016/j.hrthm.2026.04.022

26. Luther V, Chiong J, James C, et al. Diffuse right coronary artery spasm occurring 45 minutes after pulsed field ablation for atrial fibrillation. Heart Rhythm. 2025;22(7):1864–1867. doi:10.1016/j.hrthm.2025.03.1978

